# Prognostic value of Coronary artery calcium score for the prediction of atherosclerotic cardiovascular disease in participants with nonalcoholic fatty liver disease: Results from the Multi-Ethnic Study of Atherosclerosis

**DOI:** 10.1101/2023.02.20.23286213

**Authors:** Keishi Ichikawa, Spencer Hansen, Venkat S. Manubolu, Leili Pourafkari, Hooman Fazlalizadeh, Jairo Aldana-Bitar, Lisa B VanWagner, Srikanth Krishnan, Matthew J. Budoff

**Affiliations:** Lundquist Institute, Harbor-UCLA Medical Center, Torrance, California; Department of Biostatistics, University of Washington, Seattle, Washington; Department of Medicine, Division of Digestive and Liver Diseases, University of Texas Southwestern Medical Center, Dallas, Texas; Department of Medicine, Divison of Cardiology, University of California Los Angeles, Westwood, California

**Keywords:** coronary artery calcium, cardiac computed tomography, atherosclerotic cardiovascular disease, risk assessment, nonalcoholic fatty liver disease

## Abstract

**Background:** Nonalcoholic fatty liver disease (NAFLD) is associated with an increased risk of atherosclerotic cardiovascular disease (ASCVD) events, thus a diagnostic approach to help identify NAFLD patients at high risk is needed. In this study, we hypothesized that coronary artery calcium (CAC) screening could help stratify the risk of ASCVD events in NAFLD patients.

**Methods:** A total of 718 NAFLD participants from Multi-Ethnic Study of Atherosclerosis (MESA) without previous cardiovascular events were followed for the occurrence of incident ASCVD. NAFLD was defined using non-enhanced computed tomography and liver/spleen attenuation ratio <1. Cox proportional hazards regression models were used to estimate hazard ratios (HR). C-statistic and net reclassification improvement were used to compare incremental contributions of CAC score when added to the clinical risk factors.

**Results:** In multivariable analyses, CAC score was found to be independently associated with incident ASCVD (HR = 1.33, 95% CI = 1.22–1.44, p < 0.001). The addition of CAC score to clinical risk factors increased the C-statistic from 0.677 to 0.739 (p < 0.001) and the net reclassification index was 0.721 (95% CI = 0.494–0.977). In subgroup analyses, the incremental prognostic value of CAC score was more significant in NAFLD participants with low/borderline- (<7.5%) and intermediate- (7.5–20%) 10–year ASCVD risk score.

**Conclusions:** The inclusion of CAC score in global risk assessment was found to significantly improve the classification of incident ASCVD events in participants with NAFLD, indicating a potential role for CAC screening in risk assessment.

**Clinical Perspective:** With the increasing prevalence of nonalcoholic fatty liver disease (NAFLD) individuals, there is an unmet need for a diagnostic approach to identify NAFLD individuals who are at higher risk for atherosclerotic cardiovascular disease (ASCVD) events. This study showed that higher coronary artery calcium (CAC) score was associated with ASCVD events during follow-up and improved the discriminative ability for future events in NAFLD individuals. Our study suggests routine CAC screening can be useful in assessing the risk of future ASCVD events. Future studies are needed to explore the therapeutic implications of CAC screening in this population.

## Introduction

Based on the rising prevalence of obesity and type 2 diabetes, nonalcoholic fatty liver disease (NAFLD) has become the most common liver disease globally. In the United States, the prevalence of NAFLD is estimated at 25% and is projected to increase to 33.5% by 2030^1,2^. More NAFLD patients die from atherosclerotic cardiovascular disease (ASCVD) than liver-related complications, and many studies have identified NAFLD as an independent risk factor for ASCVD events beyond its associated comorbidities ^3 4^. As the global prevalence of NAFLD increases and healthcare costs rise, the prevention of ASCVD events in NAFLD patients has become a critical public health measure.. The 10–year ASCVD risk score has been widely adopted in clinical practice to estimate the risk in the adults without ASCVD. Intermediate and high 10–year ASCVD score (≥7.5%) can also be used to risk stratify NAFLD patients ^5^. while the other recent study has reported the possibility of underestimating the absolute risk in NAFLD patients^6^. Thus, there is an unmet need for data to guide clinicians in how to risk stratify NAFLD patients regarding future risk for ASCVD.

Coronary artery calcification (CAC) score determined by non-enhanced computed tomography (CT) is a simple, quick, and easy test that is available to assess the presence and extent of coronary atherosclerosis. It is well established that CAC score is a powerful predictor of ASCVD events in various populations^7^. Furthermore, previous studies have demonstrated that CAC score provided additional information beyond clinical risk scores ^8 9^. However, despite the potential for using CAC score to identify higher risk for ASCVD events among NAFLD patients, there have been no studies to date evaluating the prognostic value of CAC score in this patient population. As a result, there is currently insufficient evidence to routinely recommend CAC screening for risk stratification in NAFLD patients.

In this study, we sought to evaluate the prognostic value of CAC score in participants with NAFLD by using the multi-ethnic study of atherosclerosis (MESA) cohort. Understanding this topic will allow for a more comprehensive approach to evaluation the risk of ASCVD risk in individuals with NAFLD.

## Materials and Methods

### Study population

We included participants from the Multi-Ethnic Study of Atherosclerosis (MESA), a prospective cohort study of 6814 men and women aged 45 to 84 years without known cardiovascular disease at the time of enrollment. The study enrolled individuals from four race/ethnicity groups (white [38.5%], African American [27.5%], Hispanic [22.1%], and Chinese [11.9%]) at 6 different sites in the United States (Baltimore, Maryland; Chicago, Illinois; Forsyth County, North Carolina; Los Angeles, California; New York, New York and St. Paul, Minnesota). from July 2000 through August 2002. The study design of MESA has been previously published ^10^. All study participants provided written informed consent, and the aggregated data was deidentified. The study was approved by the institutional review board of each site.

Figure 1 shows the flow diagram of the study design. A total of 4389 participants had an adequate non-enhanced CT imaging to diagnose fatty liver. Of them, we excluded 226 participants with a history of heavy alcohol use (>14 drinks/week for men and >7 drinks/week for women), known liver disease, or oral corticosteroid or amiodarone use. Additionally, 30 participants were excluded due to missing variables for alcohol use, liver disease, and corticosteroid/amiodarone use. After these exclusion, 4133 participants remained, and 718 participants meeting criteria for NAFLD were included in this study. Among these 718 participants, five were missing status on ASCVD, two were missing total cholesterol and high-density lipoprotein (HDL) cholesterol values, three were missing smoking status, and one was missing data on diabetes status.

**Figure 1.**
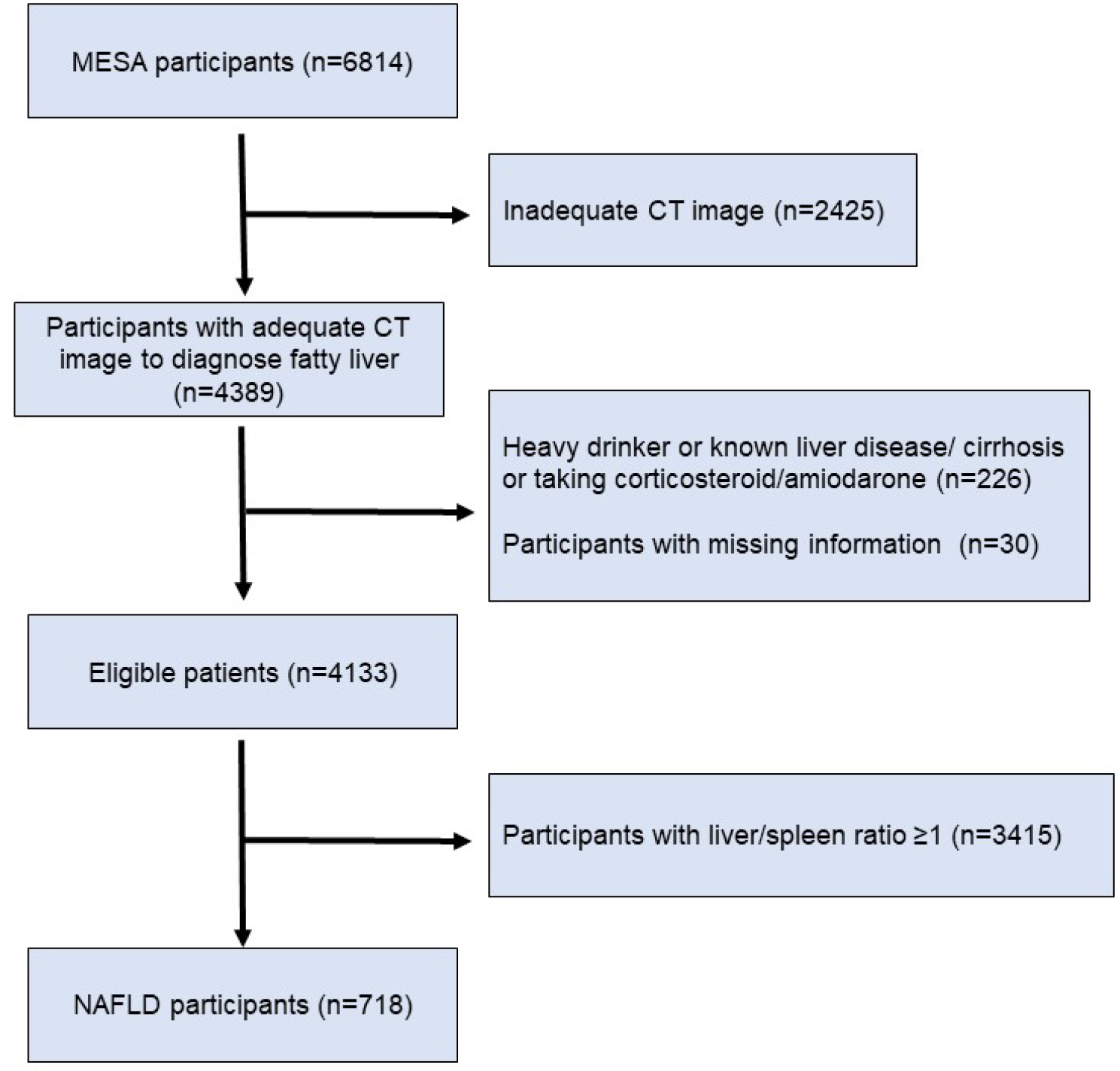
Flowchart showing the study design.

### Assessment of risk factors

As part of the baseline examination, staff at each of the six centers collected information about cardiovascular risk factors. Medical history, anthropometric measurements and laboratory data were collected during the first examination of individuals from the MESA cohort (July 2000 to August 2002). Information about age, gender, ethnicity and medical history was obtained through questionnaires. Ever smoker was defined as current and former smoker. Current smoker was defined as individuals with cigarette smoking in the last 30 days, whereas former smoker was defined as an individual who had not smoked in the last 30 days but had smoked ≥100 cigarettes in his/her lifetime. Diabetes was defined as fasting glucose of ≥7.0 mmol/l (126 mg/dl) or use of hypoglycemic drugs. Resting blood pressure was measured with the mean of the last 2 of 3 blood pressure measurements used. Total cholesterol, HDL cholesterol and triglyceride levels from blood samples obtained after a 12-h fast were measured at the Collaborative Studies Clinical Laboratory at Fairview-University Medical Center (Minneapolis, Minnesota). The 10-year ASCVD risk score was calculated from the pooled cohort equation ^11^. In this study, we classified the study population into three groups based on their risk level: low/borderline (<7.5%), intermediate (7.5–20%), and high-risk (>20%). This classification was based on a prior study ^5^.

### Assessment of NAFLD

Details of the liver fat measurement within MESA have been previously reported ^12^. Baseline cardiac CT scans were utilized to measure hepatic and splenic attenuation values (Hounsfield units) using a region of interest of ≥100 mm^2^ in area. Two regions in the right hepatic lobe and one in the spleen were measured. The liver/spleen attenuation ratio was calculated using the mean of the hepatic measurements divided by the splenic attenuation value. Hepatic steatosis was defined as liver/spleen attenuation ratio <1. NAFLD was defined after self-reported exclusion of secondary causes of liver fat (e.g., alcohol, medications) listed previously.

### Assessment of CAC score

The CAC score was measured using cardiac-gated electron-beam or multidetector CT, and details of scanning acquisition have been published previously ^13^. The CAC scores were measured with either a cardiac-gated electron-beam CT scanner (Chicago, Los Angeles, New York) or a multidetector CT (Baltimore, Forsyth County, St. Paul) at baseline and adjusted using a standard calcium phantom for calibration. All images were interpreted at the MESA CT reading center (Lundquist Research Institute, Torrance, California). We transformed the CAC score by taking the logarithm of CAC score+1 in order to maintain the normality of CAC measures. The CAC scores were also categorized as 0, 1–100, and > 100 ^14^.

### Outcome data

Participants were followed from baseline examination (2000–2002) through 2019. They were contacted by telephone every 9–12 months to inquire about interim hospital admissions, cardiovascular outpatient diagnoses, and deaths. To verify self-reported diagnoses, information was collected from death certificates and medical records for all hospitalizations and outpatient cardiovascular diagnoses. Detailed description of follow-up of MESA participants is available online (www.mesa-nhlbi.org). ASCVD events were defined as definite or probable myocardial infarction, definite or probable angina followed by coronary revascularization, definite angina not followed by coronary revascularization, resuscitated cardiac arrest, fatal or non-fatal stroke (not transient ischemic attack), other atherosclerotic death, and other cardiovascular death. A detailed description of the adjudication process has been published^10^.

### Statistical Analysis

Continuous variables are expressed as mean ± standard deviation or median with interquartile range. Dichotomous variables are expressed as numbers (proportion). Differences in continuous variables between the two groups were analyzed by the paired Student’s t-test and the Mann–Whitney U-test as appropriate. Categorical data were compared by chi-squared analysis or Fisher’s exact test. Cumulative survival estimates were calculated using the Kaplan-Meier method and compared with the log-rank test. The influence of CAC score on the occurrence of ASCVD was evaluated using the Cox proportional hazard analysis, and the results were reported as the hazard ratio (HR) with 95% confidence interval (CI). The incidence rate was expressed as the number of events per 1,000 patient-years. To determine the independent association between CAC score and ASCVD events, multivariate analysis was performed adjusting for clinical risk factors (age, sex, race/ethnicity, smoking, body mass index, total cholesterol, HDL cholesterol, lipid lowering medication use, hypertensive medication use, and diabetes). By adding the CAC score in a baseline model consisting of clinical risk factors, we assessed the discrimination and risk classification for incident ASCVD by (1) comparing the Harrell’s C-statistic (area under the curve [AUC]) of the two nested models and (2) calculating continuous net reclassification index^15^. Net reclassification index was calculated as follows = [Prob (being correctly upward reclassified/event) – Prob (being incorrectly downward reclassified/event)] + [Prob (being correctly downward reclassified /nonevent) – Prob (being incorrectly classified to an upward category/ nonevent)]. As a subgroup analysis, Cox proportional hazard model and AUC were computed for ASCVD events. All reported p values were two-sided and p<0.05 was considered statistically significant. Statistical analyses were performed using the R statistical packages *survivalROC* and *nricens* (version 4.2.1; R Foundation for Statistical Computing, Vienna, Austria).

## Results

### Patient characteristics

The mean age of the participants was 61 years old, 47% were men, 35% identified as White, 32% identified as Black, 23% identified as Hispanic, and 9% identified as Chinese American. Table. 1 shows the main clinical and laboratory baseline characteristics of the study participants with or without ASCVD events. Participants with ASCVD events were older, more likely to be male, and had higher prevalence of a high 10-year ASCVD score. There were no differences in race/ethnicity, the prevalence of diabetes, or the use of lipid lowering or hypertensive medications. However, patients with ASCVD events had higher CAC score.

### Association of CAC score and ASCVD events in NAFLD participants

During a median follow-up of 17.2 years, 138 ASCVD events occurred (coronary heart disease death [n=10], myocardial infarction [n=28], resuscitated cardiac arrest [n=3], definite angina [n=38], probable angina followed by coronary revascularization [n=4], stroke [n=45], other cardiovascular death [n=10]) were recorded. Figure. 2A shows the Kaplan-Meier curves stratified by baseline CAC score. NAFLD participants with higher CAC score at baseline had a greater event rate (Supplemental. Figure). Figure 2B shows the Kaplan-Meier curves stratified by 10-year ASCVD risk. Compared with low/borderline-risk group, intermediate- and high-risk group were significantly associated with higher event rates. As shown in Table 2, univariate Cox regression analysis identified that log(CACS+1) was significantly associated with ASCVD events (HR, 95% CI 1.38, 1.29–1.48, p < 0.001). Furthermore, multivariate Cox regression analysis identified that log(CACS+1) was independently associated with ASCVD events after adjustment for clinical risk factors (HR, 95% CI 1.33, 1.22–1.44, p < 0.001). By adding log(CACS+1) to clinical risk factors, the Harrell’s C-statistic increased from 0.677 to 0.739 (p < 0.001) (Table.3). The net reclassification index achieved by adding log(CACS+1) to clinical risk factors was 0.721 (95% CI, 0.494–0.977). Thus, the discrimination and reclassification for incident ASCVD were improved when CAC score was added to clinical risk factors.

**Table 1.** Baseline clinical and laboratory characteristics of the study population

|  | All<br>n=718<sup>^</sup> | ASCVD (+)<br>n=138 | ASCVD (-)<br>n=575 | p-value* |
| --- | --- | --- | --- | --- |
| Age, years | 60.9±9.6 | 64.6±9.0 | 60.1±9.5 | <0.001 |
| Male sex, n (%) | 335 (47) | 78 (57) | 254 (44) | 0.012 |
| Body mass index, kg/m<sup>2</sup> | 31.1±5.4 | 30.7±4.8 | 31.3±5.5 | 0.266 |
| Race/ethnicity |  |  |  | 0.899 |
| White, n (%) | 252 (35) | 52 (38) | 199 (35) |  |
| Chinese American, n (%) | 74 (9) | 14 (10) | 58 (10) |  |
| Black, n (%) | 137 (32) | 24 (17) | 112 (19) |  |
| Hispanic, n (%) | 255 (23) | 48 (35) | 206 (36) |  |
| Diabetes, n (%) | 157 (22) | 37 (27) | 117 (20) | 0.123 |
| Ever smoker, n (%) | 332 (46) | 71 (51) | 259 (45) | 0.208 |
| Lipid lowering medications, n (%) | 125 (17) | 30 (22) | 94 (16) | 0.169 |
| Hypertensive medications, n (%) | 318 (44) | 71 (51) | 246 (43) | 0.081 |
| Total cholesterol, mg/dl | 195±39 | 191±35 | 196±40 | 0.121 |
| HDL cholesterol, mg/dl | 45±12 | 44±14 | 46±12 | 0.388 |
| Systolic blood pressure, mmHg | 127±21 | 133±21 | 129±21 | 0.046 |
| 10-year ASCVD Risk |  |  |  | <0.001 |
| Low/borderline -risk, n (%) | 299 (42) | 31 (22) | 265 (46) |  |
| Intermediate-risk, n (%) | 260 (36) | 57 (41) | 201 (35) |  |
| High-risk, n (%) | 154 (21) | 50 (36) | 104 (18) |  |
| CAC score, HU | [0, 77.2] | [7.6, 299.3] | [0, 36.8] | <0.001 |
| CAC category |  |  |  | <0.001 |
| 0, n (%) | 336 (47) | 27 (20) | 306 (53) |  |
| 1-100, n (%) | 220 (31) | 45 (33) | 174 (30) |  |
| >100, n (%) | 162 (23) | 66 (48) | 95 (17) |  |

Data are presented as mean ± standard deviation, number (%), or median [25<sup>th</sup>, 75<sup>th</sup> percentile].
<sup>^</sup>5 participant's ASCVD status is missing (immediately loss-to-follow up)
\*Comparison between ASCVD (+) and ASCVD (-)
ASCVD, atherosclerotic cardiovascular disease; CAC, coronary artery calcium; HDL, high-density lipoprotein; HU, Hounsfield U.

**Table 2.**
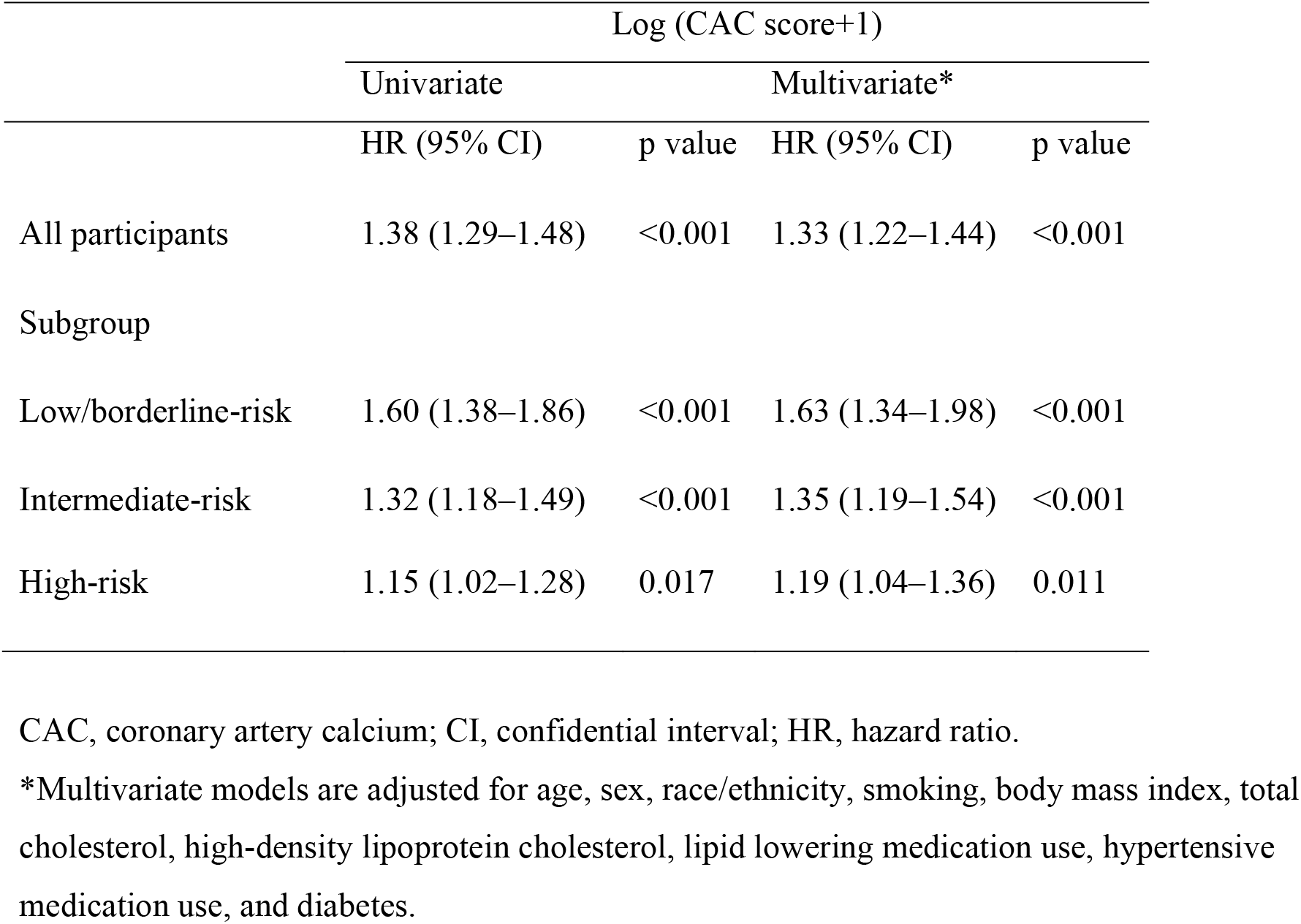
The association between CAC score and atherosclerotic cardiovascular disease events

|  | Log (CAC score+1) |  |  |  |
| --- | --- | --- | --- | --- |
|  | Univariate |  | Multivariate* |  |
|  | HR (95% CI) | p value | HR (95% CI) | p value |
| All participants | 1.38 (1.29–1.48) | <0.001 | 1.33 (1.22–1.44) | <0.001 |
| Subgroup |  |  |  |  |
| Low/borderline-risk | 1.60 (1.38–1.86) | <0.001 | 1.63 (1.34–1.98) | <0.001 |
| Intermediate-risk | 1.32 (1.18–1.49) | <0.001 | 1.35 (1.19–1.54) | <0.001 |
| High-risk | 1.15 (1.02–1.28) | 0.017 | 1.19 (1.04–1.36) | 0.011 |

CAC, coronary artery calcium; CI, confidential interval; HR, hazard ratio.
\*Multivariate models are adjusted for age, sex, race/ethnicity, smoking, body mass index, total cholesterol, high-density lipoprotein cholesterol, lipid lowering medication use, hypertensive medication use, and diabetes.

**Table 3.** Discrimination and reclassification improvement for ASCVD prediction with the addition of CAC score to clinical risk factors

|  | Harrell's C-statistic |  |  | Net reclassification index [95%CI] | Event Net reclassification index [95%CI] | Non-Event Net reclassification index [95%CI] |
| --- | --- | --- | --- | --- | --- | --- |
|  | Model 1* | Model 1+ Log(CAC score+1) | p value |  |  |  |
| All participants | 0.677 | 0.739 | <0.001 | 0.721 [0.494, 0.977] | 0.406 [0.241, 0.608] | 0.315 [0.210, 0.410] |
| Subgroup |  |  |  |  |  |  |
| Low/borderline-risk | 0.709 | 0.812 | 0.023 |  |  |  |
| Intermediate-risk | 0.593 | 0.691 | 0.009 |  |  |  |
| High-risk | 0.636 | 0.632 | 0.540 |  |  |  |

CAC, coronary artery calcium; CI, confidential interval.
\*Model 1 include age, sex, race/ethnicity, smoking, body mass index, total cholesterol, high-density lipoprotein cholesterol, lipid lowering medication use, hypertensive medication use, and diabetes.

**Figure 2.**
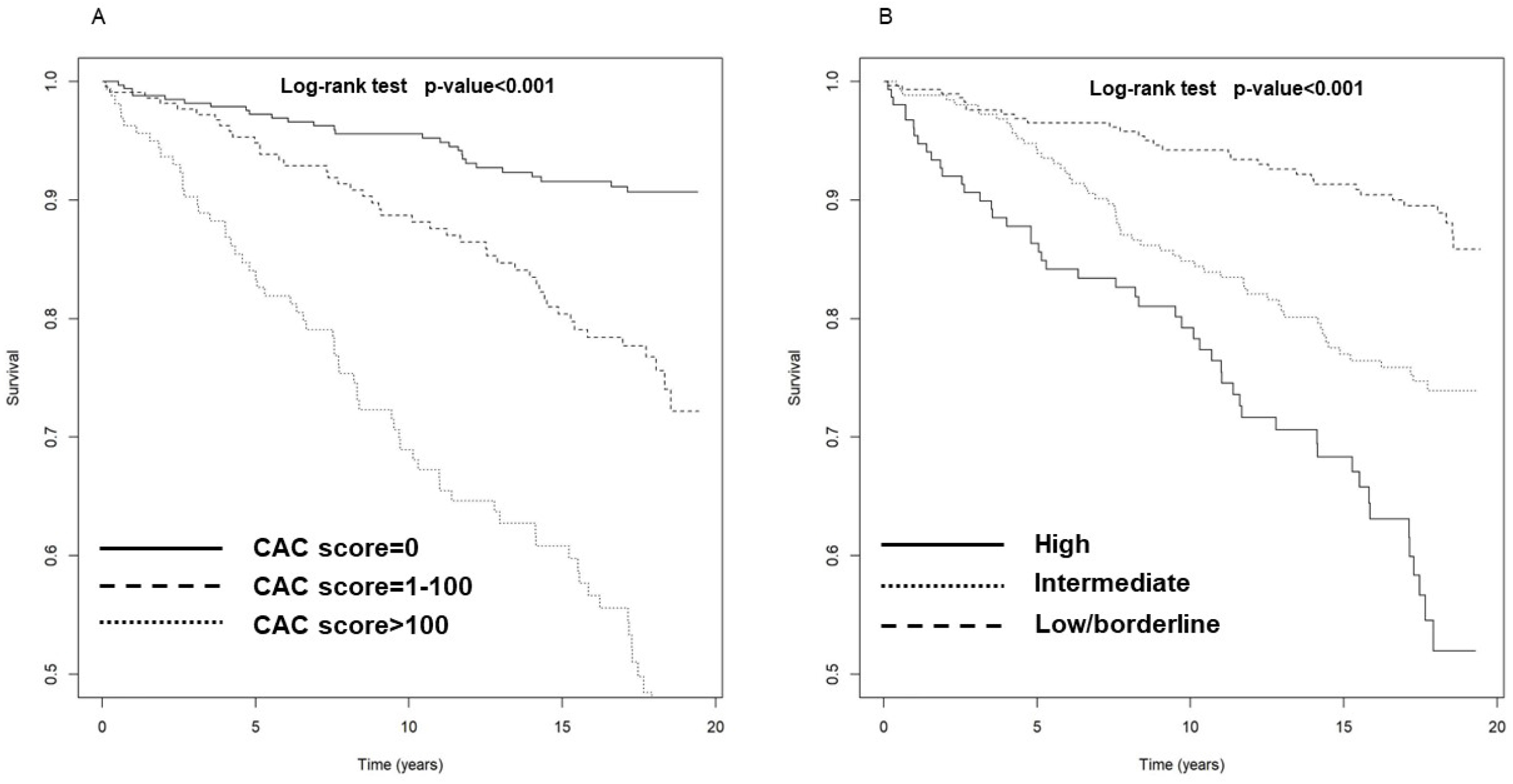
Kaplan-Meier analyses of event-free survival for ASCVD stratified by (A) CAC score or (B) 10-year ASCVD score ASCVD, atherosclerotic cardiovascular disease; CAC, coronary artery calcium; NAFLD.

**Figure 3.**
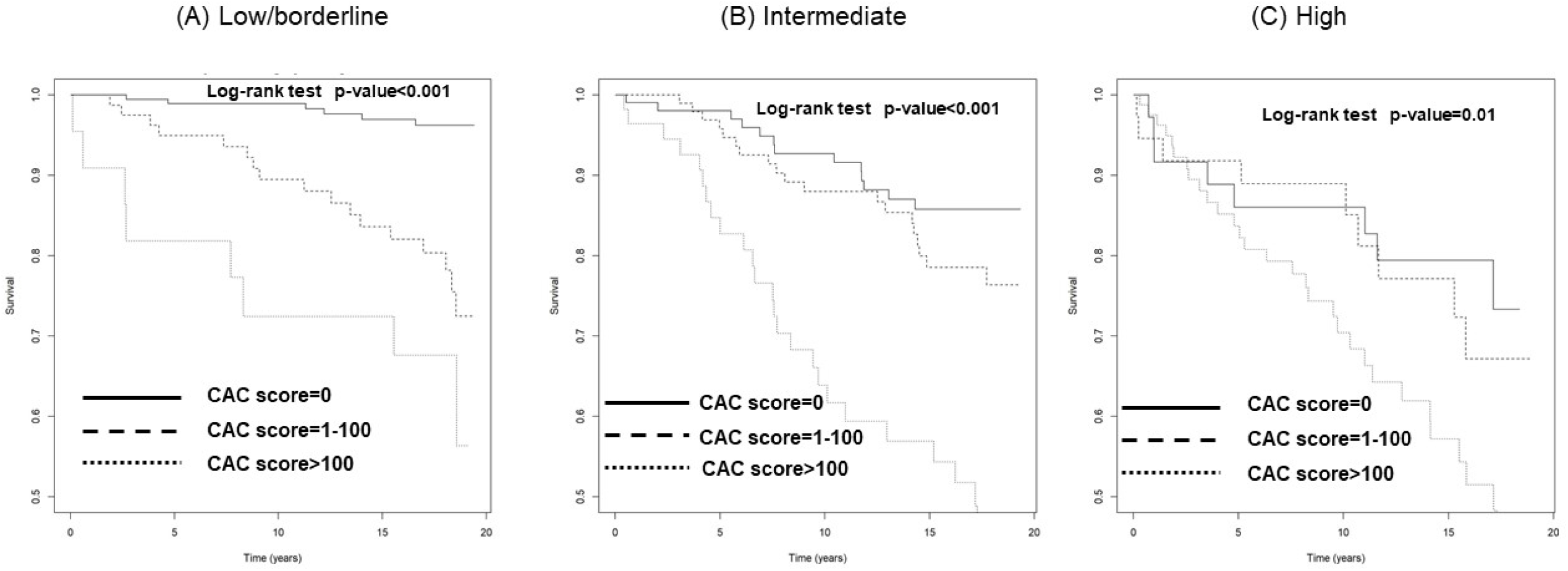
Impact of CAC score on ASCVD in NAFLD participants stratified by 10-year ASCVD risk. Participants were classified according to the following categories: 10-year ASCVD score (A) low/borderline (<7.5%), (B) intermediate (7.5–20%), (C) high (>20%). ASCVD, atherosclerotic cardiovascular disease; CAC, coronary artery calcium; NAFLD, nonalcoholic fatty liver disease.

### Association of CAC score and ASCVD events in NAFLD participants stratified by 10-year ASCVD risk

The additional predictive values of CAC score for ASCVD events were further analyzed when NAFLD participants were stratified by 10-year ASCVD risk score (low/borderline-risk group [n= 299], intermediate-risk group [n= 260], high-risk group [n= 154]). As shown in Supplemental Figure and Figure3A-3C, participants with a CAC score >100 had greater event rates in all groups stratified by 10-year ASCVD risk score. Of note, participants with CAC score of 0 had very low ASCVD event rates in low/borderline-risk group with 2.01 events per 1,000 patient-years, whereas those in intermediate- and high-risk groups had relatively high event rates with 8.59 events per 1,000 patient-years and 16.98 events per 1,000 patient-years, respectively (Supplemental. Figure). In multivariate Cox regression analysis, log(CAC score+1) was independently associated with ASCVD events after adjustment for clinical risk factors in all groups (Table. 2). By adding log(CAC score+1) to clinical risk factors, the Harrell’s C-statistic changed from 0.709 to 0.812 (p=0.023) in low/borderline-risk group, 0.593 to 0.691 (p=0.009) in intermediate-risk group, and 0.636 to 0.632 (p=0.540) in high-risk group (Table. 3). Thus, the improvements of the event prediction by addition of CAC score were more significant in low/borderline- and intermediate-risk participants.

## Discussion

To our knowledge, this is the first study to investigate the prognostic value of CAC score in NAFLD participants. Three main findings emerged from our study. Firstly, CAC scores were found to be an independent predictor of incident ASCVD events in participants with NAFLD. Secondly, participants with CAC score of 0 had very low ASCVD event rates in low/borderline-risk score group, while those in intermediate- and high-risk score groups had relatively high event rates. Thirdly, including CAC scores in addition to clinical risk factors significantly improved the ability to predict incident ASCVD events, with the greatest improvement seen in the low/borderline- and intermediate-risk score groups.

A strong association between NAFLD and increased risk of ASCVD events and mortality has been well-known ^4,16^, and there is an unmet need for a diagnostic approach to identify NAFLD patients who are at higher risk and will benefit from prevention therapy. Measuring CAC score has been shown to have strong prognostic value for ASCVD events in various populations, including type 2 diabetes individuals ^17^, a group of patients where NAFLD is extremely prevalent ^18^. Although several cross-sectional studies have found an independent association between NAFLD and the presence of CAC^19,20^, there have been no longitudinal studies showing the role of CAC score in assessing ASCVD risk in NAFLD patients. In this context, our study demonstrated the value of CAC screening in identifying NAFLD individuals at higher risk for ASCVD events. These results suggest that measuring CAC score may be helpful in determining the appropriate strategy for preventive therapies. However, future prospective studies are needed to investigate the therapeutic implications of CAC screening in NAFLD individuals.

A CAC score of 0 is established as a powerful negative risk marker for ASCVD events over a 10–15 period ^21,22^. According to current guidelines, intermediate-risk individuals with a CAC score of 0 can avoid statin therapy ^23^. However, our study demonstrated that NAFLD participants with a CAC score of 0, particularly those in the intermediate-and high-risk categories, had relatively higher event rates. Our results suggest that using a CAC score of 0 to downgrade estimated risk is not appropriate for individuals with NAFLD, as with other conditions such as diabetes, a strong family history of premature coronary heart disease, and current smoking. The mechanisms involved in the higher risk of despite a CAC score of 0 in NAFLD participants are partly explained by the association between NAFLD and plaque progression. NAFLD has been reported as an independent predictor of CAC progression^24^, indicating the presence of NAFLD may promote the conversion from a CAC score of 0 to a CAC score greater than 0. However, it should be noted that this claim is based on the observational nature of this study, and future research is needed to confirm this relationship.

The recent study by Golabi et al. showed that patients with NAFLD who have an elevated ASCVD risk score ≥ 7.5% have a higher risk of cardiovascular mortality, confirming the usefulness of such scoring systems in identifying patients at risk in this population^5^. On the other hand, Henson et al. recently found that the scoring system used in their study may underestimate the absolute risk in NAFLD patients ^6^. In our study, we also observed that the CAC score had incremental prognostic value in NAFLD participants with a lower 10-year ASCVD risk (< 7.5%), demonstrating the complementary role of CAC score in these participants. Some previous studies have also shown that CAC screening in lower risk individuals leads to efficient risk stratification. Kavousi et al. in their meta-analysis of CAC screening in women with 10-year ASCVD risk <7.5% found an increase in incident ASCVD for those with any CAC score compared to those with a score of 0 ^25^. NAFLD participants tend to be younger compared with non-NAFLD participants in the previous MESA study ^20^. Since 10-year ASCVD risk is heavily driven by age, these individuals may be estimated to have lower risk despite the presence of risk factors and an elevated lifetime risk of ASCVD^26^. Our results recommend the use of CAC screening in individuals with NAFLD, even if they are estimated as lower risk score.

## Study Limitations

This study had some limitations. First, NAFLD was diagnosed using CT which has limited specificity compared to using histology or magnetic resonance imaging^27^. Second, the histological severity of NAFLD was not available in this study. However, CT is a useful tool for diagnosing NAFLD without the complications associated with invasive methods. In addition, fibrosis markers such as the NAFLD fibrosis score and fibrosis-4 score were also not available. Previous studies have shown that fibrosis scores are associated with an increased risk of ASCVD events ^28,29^. Future studies should explore the incremental value of CAC score over these markers. Third, our risk estimates only pertain to the population studied and may not be generalizable to other populations. Finally, our risk estimates relate to baseline measures only and do not consider the possible effects of longitudinal changes in risk factors.

## Conclusions

CAC score improved the model fit and demonstrated a discriminative ability for predicting ASCVD events in NAFLD participants better than clinical risk factors. This suggests routine CAC screening can be useful in assessing the risk of future ASCVD events in these individuals. However, CAC score of 0 was associated with relatively high event rates in intermediate- and high-risk score groups, indicating that caution should be exercised when using CAC scores to lower estimated risk. Further investigation is needed to explore the therapeutic implications of CAC screening in NAFLD individuals.

## Data Availability

The MESA data are available to qualifying investigators directly from the study and also through dbGaP (http://www.ncbi.nlm.nih.gov/gap) and BioLINCC (https://biolincc.nhlbi.nih.gov).

## Figure legends

**Supplemental Figure.** Incidence of ASCVD per 1000 Person-years in all NAFLD participants and stratified by 10-year ASCVD risk score.

ASCVD, atherosclerotic cardiovascular disease; CAC, coronary artery calcium; NAFLD, nonalcoholic fatty liver disease.

